# Patient preferences for portable versus table-mounted visual field devices in rural Alabama: a mixed methods study within a telemedicine setting

**DOI:** 10.64898/2026.04.23.26351565

**Authors:** Ellen K. Antwi-Adjei, Sourav Datta, Christopher A. Girkin, Cynthia Owsley, Lindsay A. Rhodes, Matthew Fifolt, Lyne Racette

**Author notes:** Corresponding author: Lyne Racette, PhD (LR).

## Abstract

**Purpose:** To evaluate patient satisfaction and preferences for portable versus table-mounted visual field (VF) devices in a rural telemedicine setting and identify influencing factors.

**Methods:** We conducted a sequential explanatory mixed methods study at three Federally Qualified Health Centers (FQHCs) within the Alabama Screening and Intervention for Glaucoma and eye Health through Telemedicine (AL-SIGHT) study. Participants completed VF testing with table-mounted Humphrey Field Analyzer (HFA), tablet-based Melbourne Rapid Fields (MRF), and virtual reality (VR)-based VisuALL perimeters. Participants rated satisfaction, comfort, ease of use, and future testing preference. Chi-square tests assessed differences in device preferences. Twelve participants completed semi-structured interviews to explore reasons underlying preferences. Qualitative data were analyzed in NVivo 14 using reflexive thematic analysis.

**Results:** Among 271 respondents (mean age 60.4 years; 62.4% women), 50.6% preferred VR-based, 35.1% tablet-based, and 14.4% table-mounted for future testing (χ² (2) = 53.52, p<0.001, Cramér’s V = 0.31). Satisfaction was highest for VR-based (56.9% very satisfied), followed by tablet-based (49.4%), and HFA (38.0%). VR-based perimeter was most frequently selected as the most comfortable (55.7%; χ² (2) = 63.33, p<0.001, V = 0.34) and easiest to use (54.6%; χ² (2) = 71.96, p<0.001, V = 0.36). Preferences did not vary significantly across demographic variables (all p>0.05). Qualitative themes identified four key drivers: comfort and physical experience, visual experience, ease of use and interaction, and psychological and motivational factors. Portability and community suitability were valued.

**Conclusion:** Rural underserved patients strongly preferred portable visual field devices, particularly VR-based, over table-mounted HFA. Comfort, ergonomic flexibility, immersive visual experience, and simplicity of interaction were central determinants of preference. Portable perimetry may enhance patient-centered glaucoma monitoring within telemedicine programs and access in resource-limited settings.

## Introduction

Vision impairment remains a major public health concern in the United States, with glaucoma representing a leading cause of irreversible blindness.[1–4] In the United States, more than four million people are living with glaucoma, and around two million adults aged ≥40 years have the most common form, open-angle glaucoma.[2] Because glaucoma is often asymptomatic in its early stages, timely detection and ongoing monitoring are essential to prevent vision loss.[5, 6] Visual field (VF) testing is central to glaucoma diagnosis and management, providing critical information on functional impairment and disease progression.[7]

Despite its clinical importance, conventional perimetry poses practical challenges. The clinical standard, the Humphrey Field Analyzer (HFA), is expensive, stationary, and requires a dedicated space, chin/forehead support, eye patch, and technician supervision.[8] These structural and procedural requirements can make testing uncomfortable and limit availability in resource-constrained settings. Patients frequently report VF testing as necessary but tiring or burdensome, and barriers such as travel distance and prolonged clinic visits may reduce adherence to follow-up care, particularly in rural and underserved communities. [9–14]

Telemedicine has emerged as a strategy to expand access to glaucoma screening and monitoring in these populations by enabling remote acquisition and specialist interpretation of ocular data.[11, 15–18] However, integrating VF testing into ophthalmic telemedicine programs remains challenging because conventional clinical standard perimeters are bulky, costly, immobile, require specialized equipment, and not easily deployed outside specialty clinics.

Portable visual field technologies, including tablet-based and head-mounted virtual reality (VR)-based perimeters, have been developed to address these limitations. These devices are lightweight, comparatively affordable, and can be operated without dark rooms, chin rests, or trial lenses, making them suitable for community and primary care settings. [8, 19–21] Studies suggest that portable devices demonstrate promising agreement with standard perimetry.[21–24] Nevertheless, little is known about how patients in medically underserved rural settings perceive these technologies relative to the clinical standard. Understanding patient satisfaction, usability, and preference is critical for designing patient-centered ophthalmic telemedicine programs and may influence long-term adherence to glaucoma monitoring.

To address this gap, we conducted a sequential explanatory mixed methods study embedded within a rural telemedicine setting to compare patient experiences with three VF devices: the HFA, the tablet-based Melbourne Rapid Fields (MRF), and the VR-based VisuALL. We aimed to (1) quantify patient satisfaction and preferences for future testing, and (2) qualitatively explore the experiential factors influencing device choice. By integrating patient perspectives into ophthalmic telemedicine care models, this study seeks to inform the implementation of portable perimetry in underserved rural settings.

## Method

### Study Design and Setting

We employed a sequential explanatory mixed methods design comprising a cross-sectional quantitative survey followed by in-depth qualitative interviews. Participants were recruited from three Federally Qualified Health Centers (FQHCs) participating in the Alabama Screening and Intervention for Glaucoma and eye Health through Telemedicine (AL-SIGHT) study—Centreville (Bibb County), Maplesville (Chilton County), and Marion (Perry County), Alabama.[11, 16] These sites serve predominantly rural communities located within or adjacent to Alabama’s Black Belt region, historically named by its rich, fertile black soil.[25] The region has predominantly African American population and longstanding socioeconomic and healthcare disparities, with limited access to specialty eye care, making it an important area for vision research and targeted intervention.[26, 27] The protocol was approved by the Institutional Review Board of the University of Alabama at Birmingham, and all procedures adhered to the tenets of the Declaration of Helsinki. Written and verbal informed consent was obtained from all participants.

### Quantitative Phase

#### Participants and Data Collection

Adult patients (≥18 years) attending telemedicine visits between November 2022 and November 2023 completed VF testing using the HFA3 (Carl Zeiss Meditec, Dublin, CA), the Melbourne Rapid Fields tablet perimeter (MRF; M&S Technologies, Niles, IL), and the VisuALL VR headset (Olleyes Inc., Summit, NJ). After completing all three tests, participants completed an interviewer-administered structured questionnaire.

Questionnaire items were derived from existing perimetry and usability literature to ensure comprehensive coverage of constructs. The instrument was reviewed by experts in glaucoma, vision science, and qualitative research (LAR, LR, MF) to assess its relevance and appropriateness. It was subsequently pilot-tested to evaluate clarity, comprehension, and flow prior to final administration, and further refinements were made accordingly.[28–31] The survey was administered by trained site research coordinators who read each question aloud to participants and recorded their responses, ensuring accessibility for individuals with limited literacy or vision challenges. Coordinators also made brief observation notes describing participants’ reactions and behaviors during testing; these contextual notes later informed the qualitative interview guide and served as prompts during the follow-up phase. Items included a 4-point Likert-type scale assessing satisfaction with each device (1 = very dissatisfied to 4 = very satisfied), and open-ended questions asking which device they would choose for a future test and why; which device was the most comfortable, and why; which device was easiest to use, and why. Demographic information (age, sex, race/ethnicity, employment, education, insurance status) and prior eye-examination history were also recorded.

#### Statistical Analysis

Descriptive statistics summarized participants’ characteristics, satisfaction ratings, comfort, ease of use, and preference by device. Overall differences in device preference distributions were evaluated using Chi-square goodness-of-fit tests. Associations between device preference (HFA, MRF, VisuALL) and demographic characteristics were assessed using Pearson Chi-square tests of independence. These subgroup analyses were considered exploratory. Because multiple demographic comparisons were conducted for each outcome, the potential for inflated Type I error was evaluated using the Benjamini-Hochberg false discovery rate (FDR) procedure. Differences were considered statistically significant at p < 0.05. Statistical analyses were performed in R (version 4.4.2).[32]

### Qualitative Phase

#### Sampling and Interviews

After completion of the quantitative phase, 56 participants who had completed both baseline and test-retest VF assessments formed the sampling pool for interviews. We used stratified purposive sampling to ensure representation of each preferred device and to capture diversity in age, sex, and study site. This phase ran from August 2025 to October 2025. Twelve participants were invited via telephone and agreed to participate. Because the VR-based VisuALL device was the most preferred in the quantitative results, six participants were purposely selected from those who identified VR as their preferred device across all three survey domains (future testing preference, comfort, and ease of use). Three participants each were selected from those who preferred the tablet-based MRF and the table-mounted HFA. The latter two device groups were included exploratorily to gain broader insight into contrasting experiences. Including all three device types provided a richer comparative understanding of patient experience.

Interviews were conducted remotely via Zoom (Zoom Video Communications, San Jose, CA) by a trained researcher (EKAA), who had completed graduate-level coursework in qualitative research methods and received faculty mentorship in semi-structured interviewing techniques. A semi-structured interview guide, informed by participants’ survey responses and field notes, was used to facilitate discussion.

During the interview, participants were shown images of each device (Fig 1) and invited to reflect on comfort, visual experience, ease of use, perceived accuracy, and overall preference. Follow-up questions were structured to facilitate direct comparison across devices within specific experiential domains (e.g., comfort, ease of use, and device features). Participants were also asked to elaborate on reasons for non-chosen devices, critical incidents that influenced their decisions, and suggestions for improving VF testing in primary care and community settings. Interviews lasted 15–20 minutes and were audio-recorded with participants’ permission. Initial transcripts were generated using Zoom’s automated transcription feature and subsequently reviewed and refined using Microsoft Word’s transcription tool. All transcripts were manually verified against the original audio recordings by the interviewer (EKAA) to ensure accuracy prior to analysis.

**Fig 1.**
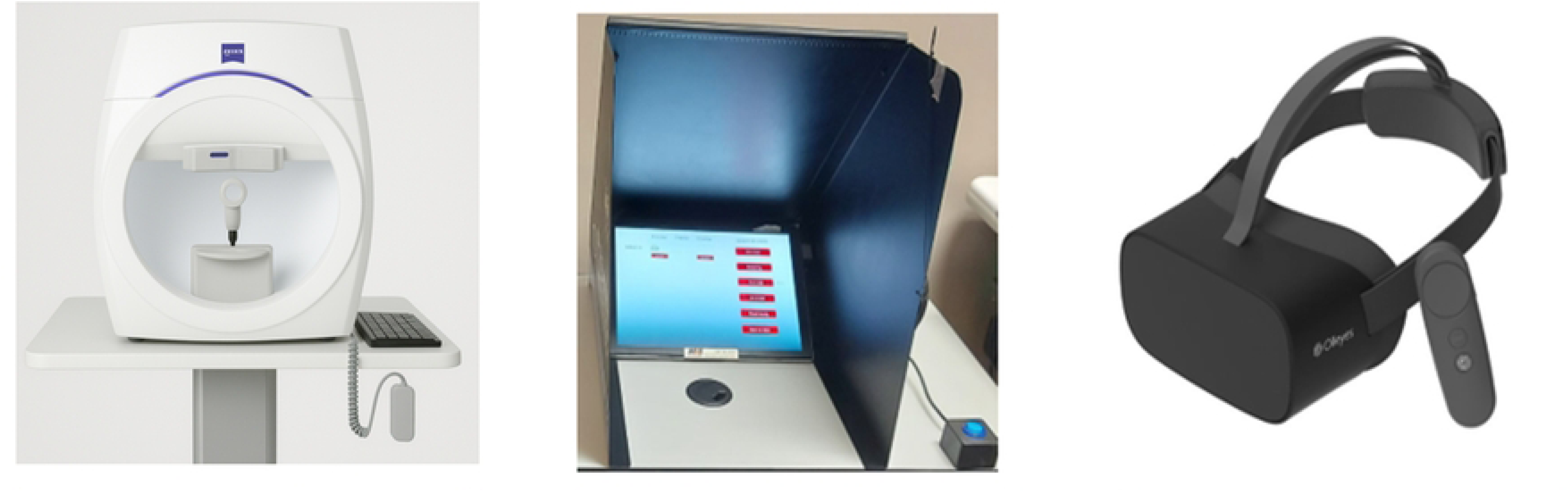
Visual field devices used in the study. Table-mounted (HFA) is the traditional VF device, while Tablet-based (MRF) and VR-based (VisuALL) represent portable VF devices.

#### Qualitative Analysis

Qualitative data were managed and analyzed in NVivo 14 using Braun and Clarke’s reflexive thematic analysis.[33] This six-phase approach involves familiarization with the data, generating initial codes, searching for potential themes, reviewing themes, defining and naming themes, and producing the report. Two members of the research team (EKAA and SD) independently immersed themselves in the interview transcripts by reading them line by line. This immersion generated descriptive, data-driven codes that were then collated into broader categories; through comparison and iterative discussion, these categories were synthesized into higher-order themes. To enhance analytical rigor, one coder (SD) was not involved in data collection, ensuring an external perspective and reducing researcher bias.

Coding discrepancies were resolved through regular meetings, and the team refined the coding framework until consensus was reached on the final themes. Inter-rater reliability, assessed with Cohen’s kappa (κ = 0.90), indicated almost perfect agreement between coders.

To ensure rigor and trustworthiness, the qualitative phase followed the principles of credibility, dependability, confirmability, and transferability as outlined by Lincoln and Guba.[34] Credibility was strengthened through coder triangulation, with both coders independently analyzing transcripts before meeting to compare interpretations and reach agreement on final themes. Dependability was established through the maintenance of a detailed audit trail in NVivo, documenting coding decisions, codebook updates, and theme development across analytic meetings.

Confirmability was supported through reflexive memoing throughout the analytic process, allowing the researchers to identify potential biases and ensure that interpretations were grounded in participant data. Transferability was enhanced by providing rich, contextualized descriptions of the study setting, participant demographics, and experiences within rural Alabama FQHCs, enabling readers to assess applicability to similar contexts. Data saturation was achieved when no new themes emerged in successive transcripts. Anonymized transcripts were used for analysis. While VR emerged as the most preferred device in the quantitative phase, thematic patterns were explored across all three devices to capture similarities and distinctions in patient experience. Collectively, these strategies ensured that the analysis was systematic, transparent, and aligned with best practices in qualitative research.

## Results

### Quantitative Findings

#### Participant Demographics

Of the 271 participants, the mean (SD) age was 60.4 (11.8) years, 62.4% were female, 66.8% White, and 30.6% African American. Nearly all participants (96.7%) had undergone a previous eye examination. Table 1 summarizes the baseline demographic and characteristics of participants.

**Table 1.** Participant baseline demographics and characteristics (N = 271)

| **Socio-demographics** | **Mean (±SD) or n (%)** |
| --- | --- |
| **Age, years** | 60.4 (±11.8) |
| **Age categories, years** |  |
| Less than 40 | 12 (4.4) |
| 40-60 | 136 (50.2) |
| 61 and above | 123 (45.4) |
| **Sex** |  |
| Female | 169 (62.4) |
| Male | 102 (37.6) |
| **Race** |  |
| White | 181 (66.8) |
| African American | 83 (30.6) |
| Other* | 7 (2.6) |
| **Ethnicity** |  |
| Hispanic | 3 (1.1) |
| Non-Hispanic | 268 (98.9) |
| **Employment†** |  |
| Employed | 81 (29.9) |
| Unemployed | 22 (8.1) |
| Retired | 82 (30.3) |
| Unable to work | 79 (29.1) |
| Other | 7 (2.6) |
| **Marital Status** |  |
| Married or domestic partnership | 143 (52.8) |
| Divorced | 63 (23.2) |
| Single | 38 (14.0) |
| Widowed | 27 (10.0) |
| **Educational Level** |  |
| Less than high school | 64 (23.6) |
| High school or equivalent | 100 (36.9) |
| Some college or associate degree | 80 (29.5) |
| College graduate | 17 (6.3) |
| Graduate School or other professional degree | 10 (3.7) |
| **Ever had an eye exam** |  |
| Yes | 262 (96.7) |
| No | 9 (3.3) |
| *There were 5 multiracial persons and 2 unknown/refused to answer<br>†Employed includes full-time, part-time, and self-employed; Other includes student, homemaker/caregiver, and other |  |

#### Satisfaction with VF Devices

Satisfaction ratings varied across devices. The VR-based VisuALL received the highest proportion of “very satisfied” responses (56.9%), followed by the tablet-based MRF (49.4%) and the table-mounted HFA (38.0%). Participants frequently commented that the portable devices were more comfortable, quicker, and easier to use than the table-mounted device. Fig 2 shows the satisfaction levels for each device.

**Fig 2.**
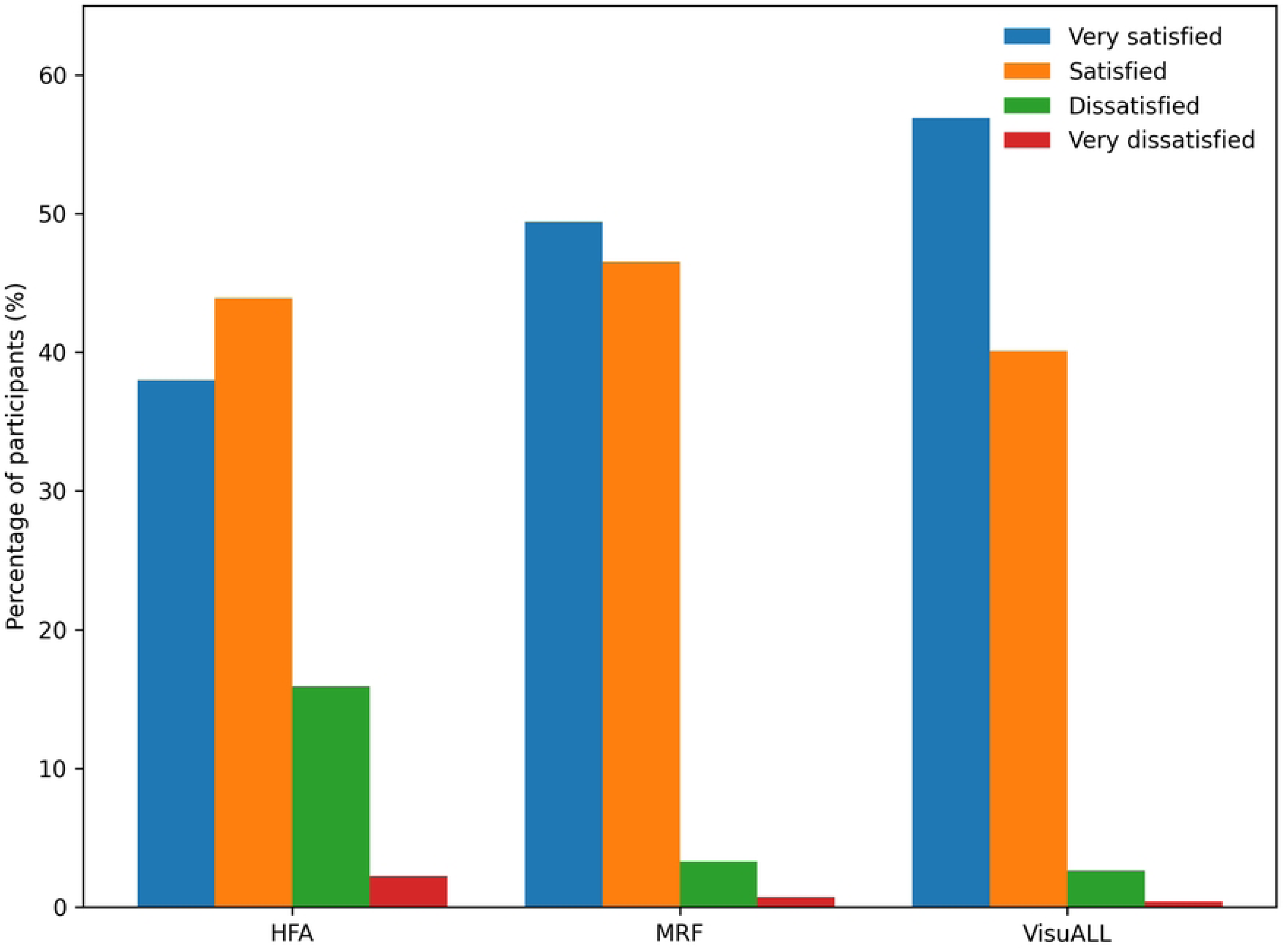
Percentage distribution of participant satisfaction ratings across visual field devices. Bars represent the proportion of participants reporting very satisfied, satisfied, dissatisfied, or very dissatisfied for the table-mounted Humphrey Field Analyzer (HFA), tablet-based Melbourne Rapid Fields (MRF), and VR-based VisuALL perimeter. Satisfaction levels are presented separately for each device to allow direct comparison across testing modalities. Percentages are based on total respondents for each item (N = 271; VR VisuALL satisfaction item n = 269)

#### Device preference for future testing

When asked which device they would choose for future VF testing, 137 participants (50.6%) selected the VR-based VisuALL, 95 (35.1%) selected the tablet-based MRF, and 39 (14.4%) selected the table-mounted HFA. A Chi-square goodness-of-fit test demonstrated that the distribution differed significantly from equal preference across devices (χ² (2) = 53.52, p < 0.001), with a moderate effect size (Cramér’s V = 0.31). Fig 3A illustrates the comparative distribution of preferred devices for future testing. Device preference for future testing was not significantly associated with age cohort, sex, race, employment status, marital status, education level, or insurance status (all p > 0.05).

**Fig 3.**
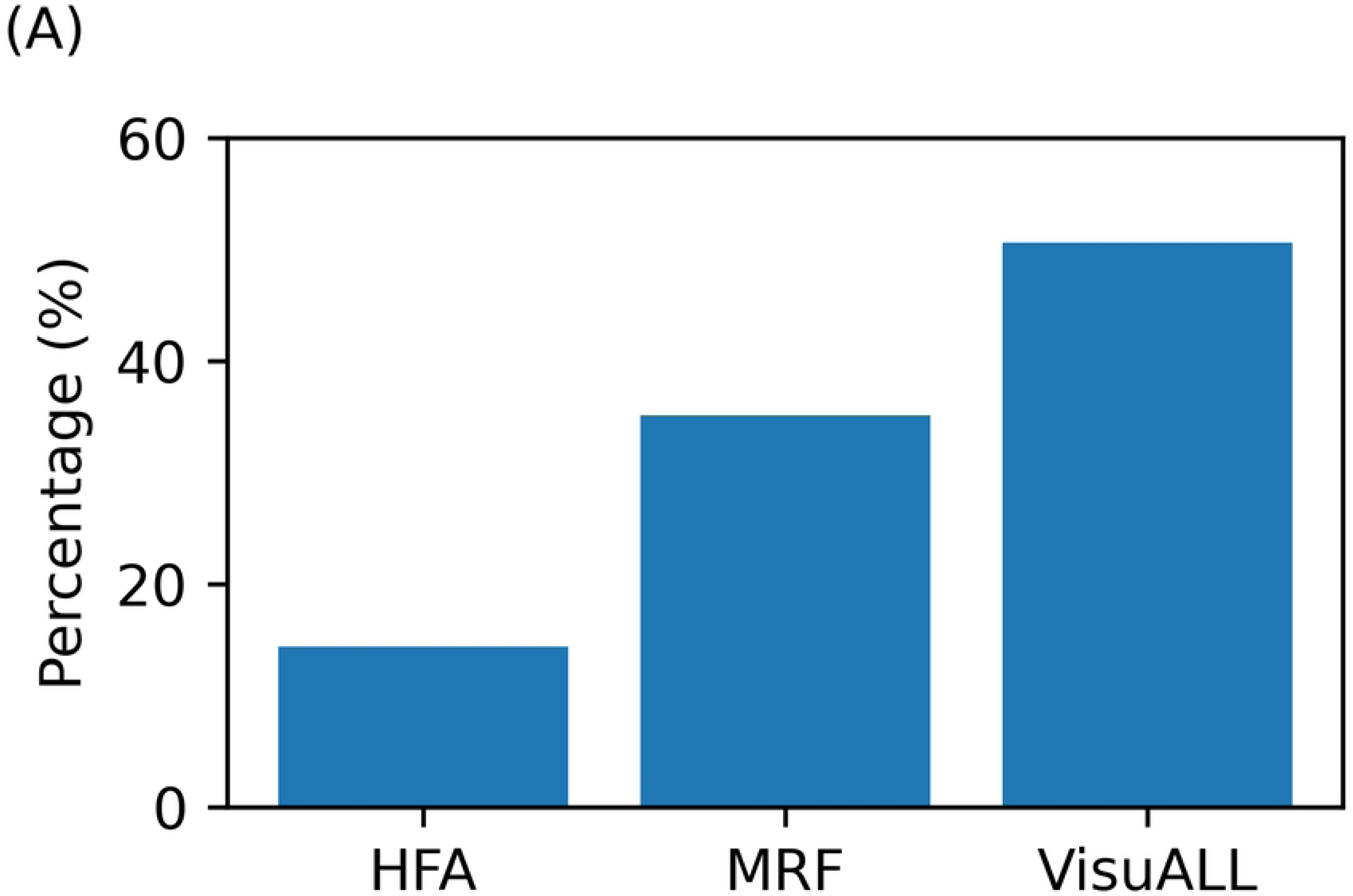
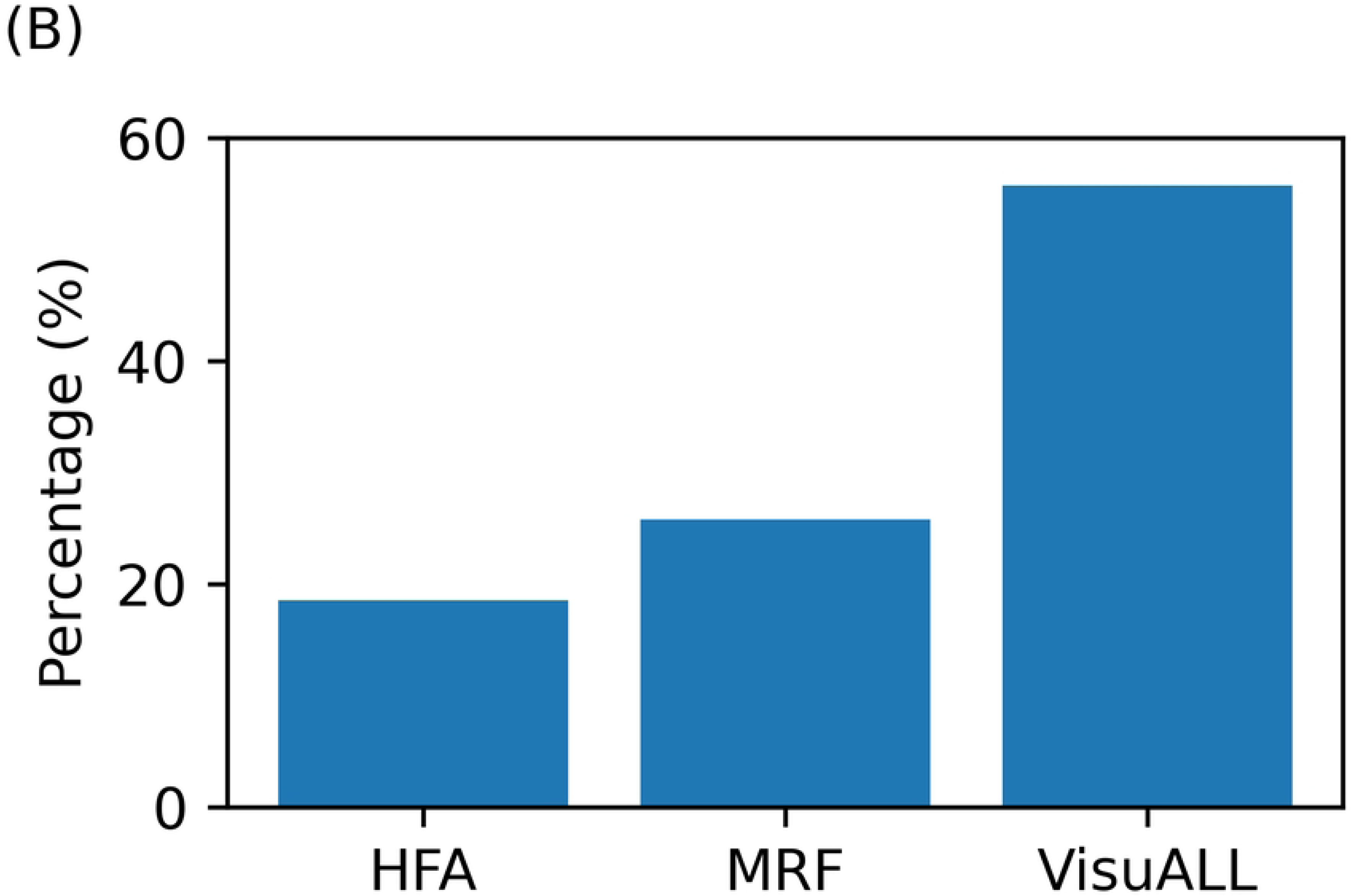
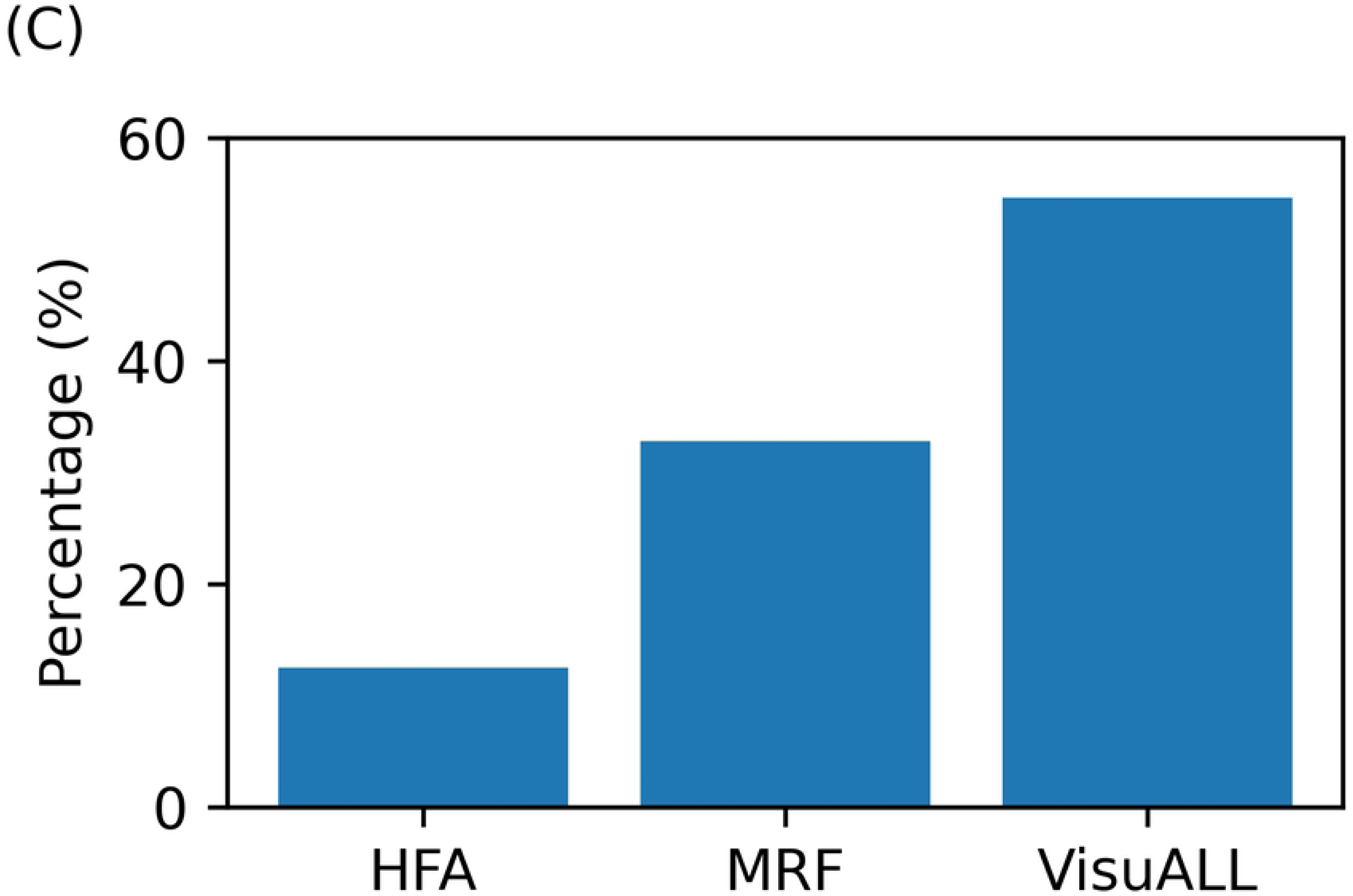
Distribution of participant-reported device preferences. [N = 271]. (A) Preferred device for the next visit; (B) Most comfortable device; (C) Easiest test to take. Bars represent the percentage of participants selecting the table-mounted Humphrey Field Analyzer (HFA), tablet-based Melbourne Rapid Fields (MRF), or VR-based VisuALL perimeter for each outcome.

#### Most comfortable device

When asked which device was most comfortable, 151 participants (55.7%) selected the VR-based VisuALL, 70 (25.8%) selected the tablet-based MRF, and 50 (18.5%) selected the table-mounted HFA. A Chi-square goodness-of-fit test demonstrated that the distribution differed significantly from equal preference across devices (χ² (2) = 63.33, p < 0.001), with a moderate effect size (Cramér’s V = 0.34). Fig 3B illustrates the comparative distribution of comfort ratings across devices. In unadjusted analyses, comfort-based device preference was significantly associated with age cohort (χ² (4) = 12.86, p = 0.01; Cramér’s V = 0.15). However, the association did not remain statistically significant after adjustment for multiple comparisons. No other demographic variables were significantly associated with comfort preference (all p > 0.05).

#### Easiest device to use

When asked which device was easiest to use, 148 participants (54.6%) selected the VR-based VisuALL, 89 (32.8%) selected the tablet-based MRF, and 34 (12.5%) selected the table-mounted HFA. A Chi-square goodness-of-fit test demonstrated that the distribution differed significantly from equal preference across devices (χ² (2) = 71.96, p < 0.001), with a moderate effect size (Cramér’s V = 0.36). Fig 3C presents the comparative distribution of ease-of-use ratings. Ease-of-use preference was not significantly associated with age cohort, sex, race, employment status, marital status, education level, or insurance status (all p > 0.05).

### Qualitative findings

Thematic analysis of 12 interviews revealed four interrelated themes explaining participants’ device preferences (Fig 4). Selected illustrative quotations for each subtheme are presented in Table 2.

**Fig 4.**
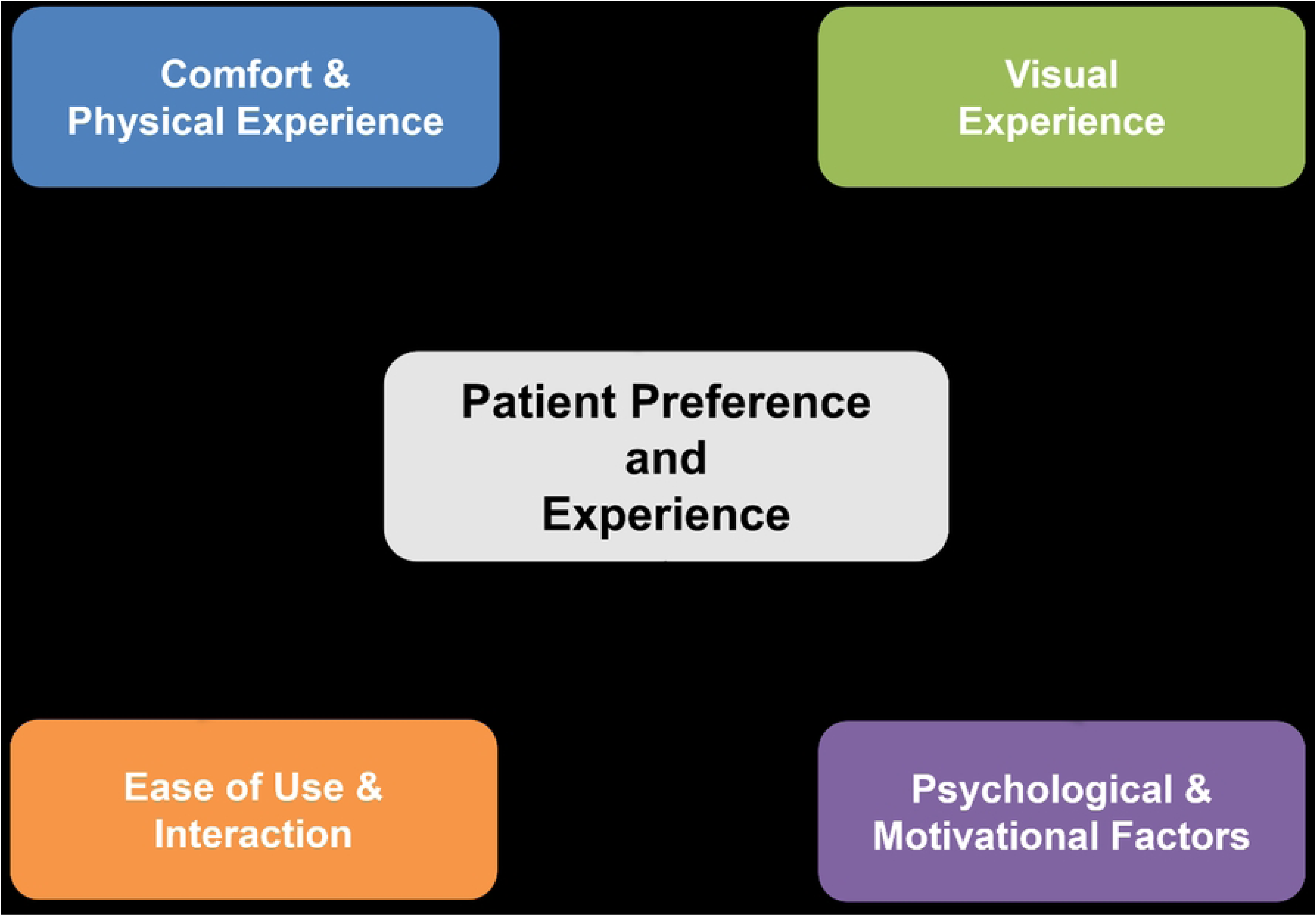
Conceptual framework of patient preference for visual-field devices. The central box represents the overarching outcome—patient preference and experience—while the four surrounding colored boxes illustrate the key themes derived from interviews. Connecting lines indicate that each theme influences the central outcome.

**Table 2.** Selected participant quotations illustrating themes and subthemes.

| **Main Themes** | **Subthemes** | **Selected Participant (P) Quotes** |
| --- | --- | --- |
| **Comfort and physical experience** | Physical comfort | *"I have a bad neck with squished vertebrae... it was hard to sit still. I could wiggle around a bit — my neck always hurts — so that was good because I could sit however I was relaxed."* (P2_VR)<br>*"The patch was very light and didn't cause me any discomfort."* (P10_Tablet)<br>*"I felt more relaxed, comfortable, and more stable with the chin and forehead rest."* (P12_Table) |
|  | Less confinement and freedom to move | *"Keeping my head braced into a bowl... too much going on... had to keep my head still. The goggles allowed me free-range motion of my head."* (P4_VR)<br>*"I felt free. I didn't feel like I was stuck... I didn't feel confined."* (P3 & P4_VR)<br>*"Resting my chin on the big bowl made me feel claustrophobic."* (P6_Tablet)<br>*"I didn't have to be still and confined in a fixed position."* (P8_Tablet) |
|  | Fit and adjustability | *"The goggles were comfortable... once adjusted, they fit well on the face and stayed in place." (P3_VR)*<br>*"The goggles were adjustable. They fit nicely and are secure. It fits around my head... they conform to my head and eyes. The tablet and table-mounted bowl were not weight-friendly for bigger people like me." (P9_VR)*<br>*"VR goggles don't sit well with me." (P6_Tablet): "There is a headrest from the shroud if needed, and I don't have to do too much to it." (P8_Tablet)*<br>*"The bowl was big enough, with head and chin well-positioned. Positioning could be adjusted to fit my height." (P12_Table)*<br>*"I like having the adjustable chinrest of the bowl, so my neck doesn't hurt." (P7_Table)* |
| **Visual experience** | Visual clarity | *"It gave me a clear picture of what I was supposed to look at." (P1_VR)*<br>*"Everything was crisp and sharp. With the other devices, I got reflections from lamps or light spots that would throw me off. (P2_VR)*<br>*"The clarity was fantastic. The screen was clear." (P6_Tablet)* |
|  |  | *"I could see the dots clearly in the background."* (P8_Tablet)<br>*"The screen is bigger, so I can see a little bit more."* (P5_Table) |
|  | Ambient light blocked and less distraction | *"With the table-mounted, you look through a screen and can still see the outside edges, but with the goggles, you don't see all that around the side."* (P1_VR) |
|  | Personal/privacy (immersion) | *"They kind of wrapped around my eyes and sealed off the sides, so I couldn't see anything else."* (P1_VR)<br>*"It felt more personal... it covered everything around me, so it felt like it was just me and the goggles."* (P3_VR)<br>*"That device that goes over the eyes gives me vertigo."* (P5_Table) |
|  | Optical comfort | *"I see better with my glasses. Both eyes were opened, but for the other two, I had to keep one eye covered, and it felt awkward."* (P3_VR)<br>*"I used my glasses. I only had to worry about one eye at a time."* (P10_Tablet)<br>*"I don't have to use my own lens. The in-built lens was helpful."* (P5_Table) |
| **Ease of use and interaction** | Simple interaction | *“Once I got them on, all I had to do was look through and do the test—it was just overall easier.” (P1_VR)*<br>*“The button was easy... just set your hand down and push.” (P6_Tablet)*<br>*“I was able to concentrate on what I was doing better.” (P12_Table)* |
|  | Ease of instructions | *“The instructions were simple.” (P3_VR)*<br>*“I like the voice of the lady speaking inside the googles. The voice instructions were clear and simple. You lose focus trying to stay still with the other devices.” (P11_VR)*<br>*“It’s very easy. You just stay focused on it, and the voice inside instructs you on when to fixate on a corner. Instructions were easy to follow.” (P8_Tablet)*<br>*“Instructions were clear and better suited to me.” (P12_Table)* |
|  | Device design suggestions and size/portability | *“They were easier for small clinics because you can store them away — no big equipment taking space.” (P2_VR)*<br>*“The setup occupied a small space and can easily fit in any room. With the bowl device, the gray dot blended into the background. I suggest a red dot on a dark background like the tablet.” (P10_Tablet)* |
|  |  | *"The bowl is big with more space for me." (P5_Table)* |
|  | Familiarity | *"My grandson has the VR game. I see him on it all the time." (P11_VR)*<br>*"The tablet was simple and easy... like working on a computer, which I am used to." (P8_Tablet)*<br>*"I've done the Humphrey test before, so I kind of know what to expect—I feel like it's reliable." (P5_Table)* |
| **Psychological and motivational factors** | Subjective preference/overall evaluation | *"Yes, definitely the goggles! Overall, it was the best one. I'm not against any of the tests; I just prefer goggles." (P1_VR)*<br>*"Ease of use was a ten out of ten compared to one or two for the others. Yes, within the first minute, it was so easier than the others." (P2_VR)*<br>*"It was a more pleasant experience than the other two. I like the experience." (P10_Tablet)*<br>*"I knew it was the right one for me; it's just comfortable." (P5_Table)* |
|  | Novelty and fun/emotional engagement | *"The goggles felt so cool. It was almost like getting ready to play a video game. They were fun and made me want to do the test." (P3_VR)* |
|  |  | *"It was a novel form of testing. Technology doesn't scare me; it attracts me."* (P10_Tablet) |
|  | Cost and practicality | *"The big table-mounted one must be expensive to buy and maintain. Goggles are cheaper and more practical for smaller places."* (P2_VR) |

### Identified Themes

#### Comfort and Physical Experience

Participants consistently emphasized comfort as a central factor influencing their device preference. Many contrasted the physical comfort provided by portable devices with the more rigid posture required by the table-mounted device. Wearing a VR headset or using a tablet allowed respondents to sit in various types of chairs, adjust their head and neck freely, and avoid leaning forward against a bowl or chinrest. Although the tablet required maintaining a fixed viewing distance from the tablet, participants perceived it as less restrictive than the table-mounted device. Several participants expressed appreciation for being able to wear their habitual glasses and avoid eye patches or trial lenses, which contributed to a more natural testing experience. In contrast, the table-mounted device’s chinrest and fixed position were frequently described as “confined,” “claustrophobic”, and “uncomfortable”, especially among individuals with neck or back pain or larger body habitus that made fixed positioning difficult. Participants noted that maintaining strict fixation and posture could cause physical strain over time. However, some respondents acknowledged that the stability provided by the table-mounted device offered a sense of structure and support, suggesting that familiarity and perceived steadiness could mitigate discomfort for certain individuals.

A related subtheme involved freedom of movement and reduced confinement. Respondents positively regarded being able to make minor head or body adjustments during testing, particularly with the VR device, which allowed slight movement without compromising the assessment. Several participants described the VR-based device as “not restricted” and noted that they “didn’t have to sit in a specific posture.” This flexibility contributed to a sense of physical ease and autonomy during testing.

Fit and adjustability also influenced comfort perceptions. Participants indicated that once the headset or device was properly adjusted, it “fit well and stayed in place.” Despite technicians optimizing seating and device positioning according to standard testing protocols, suggestions for ergonomic improvement were primarily directed toward the table-mounted device, including lowering the chair or adjusting table height to improve alignment and reduce physical strain.

In summary, comfort substantially influenced device preference. Physical comfort, less confinement and freedom to move, and fit and adjustment were the main subthemes. Participants generally favored portable devices, particularly VR-based, because they reduced physical strain, minimized confinement, eliminated the need for chinrests, and allowed a more natural posture during testing. These impressions indicate that VR-based perimetry provides a portable, immersive testing environment that may enhance comfort while minimizing external distractions.

#### Visual experience

The perceived quality of the visual environment substantially influenced device preference. Participants frequently compared the clarity of stimuli presentation across devices and described how ambient light and peripheral distractions affected their ability to focus during testing.

Clear target visibility emerged as a central component of visual experience. Both the VR and the tablet were commonly described as providing “a clear picture” and making it easy to identify stimuli. While some participants preferred the familiar optics and large bowl of the table-mounted, many noted that the portable devices offered comparable clarity. For several respondents, clarity was linked not only to the sharpness of stimuli but also to how visually contained and structured the testing environment felt.

Blocking ambient light and minimizing distractions were particularly salient advantages of the VR headset. Participants explained that the headset “sealed off the sides,” allowing them to focus exclusively on the fixation target and light stimuli without peripheral interference. As one participant stated, “you didn’t have to see things to the side… everything was right there.” This immersive visual containment was perceived as enhancing concentration and reducing external distraction. The immersive quality of the VR-based device also contributed to a sense of personal space and privacy. Several participants described feeling “isolated” or “wrapped around the eyes,” which they interpreted positively as promoting focus. Some likened the experience to a video game, suggesting that immersion made the test more engaging rather than intimidating.

Optical comfort further shaped visual experience. Participants expressed appreciation for being able to wear their habitual glasses and avoid eye patches or trial lenses, particularly with the VR headset. Binocular viewing was perceived as more natural, and some participants expressed dislike for monocular testing with an eye patch. While participants who valued broader peripheral awareness occasionally preferred the more open configuration of the tablet or table-mounted, the controlled visual environment of the VR was generally regarded as beneficial for concentration.

In summary, portable devices, particularly VR-based, were perceived as creating a visually contained and focused testing environment that enhanced clarity and engagement. Visual clarity, ambient light blocked and less distraction, immersion, and optical comfort were the main subthemes.

#### Ease of use and interaction

Simplicity and intuitive interaction were major factors influencing device preference. Participants evaluated how straightforward each device was to operate, the clarity of instructions provided, and the practicality of device design. Many respondents described VR and tablet as easy to use once positioned, requiring only a simple button press when a light was seen. Participants emphasized that the interaction felt intuitive and did not demand complex technological skills. One participant noted, “I’m not a big computer person, so anything simpler is better for me.” In contrast, although some participants were accustomed to the table-mounted, they observed that it required additional steps, including chin positioning and eye patching, which increased procedural complexity.

Instructional clarity also contributed to perceived ease. The VR and tablet devices were frequently described as requiring minimal explanation, with instructions characterized as “simple” and “easy.” The presence of guided prompts, particularly in VR, further reduced cognitive burden. By comparison, participants perceived the table-mounted setup as more involved due to alignment requirements and patching procedures.

Device design and portability were additional considerations. Participants remarked that VR-based and tablet-based portable devices required minimal space and could be used in various settings beyond traditional examination rooms. Several noted that these devices were compact and easy to store. Suggestions for ergonomic improvements were primarily directed toward the table-mounted device, including adjustments to chair height or button placement.

Familiarity also influenced perceptions of ease. Some participants preferred the table-mounted because they had used it previously and trusted its results; familiarity with the chinrest and response button reduced uncertainty and anxiety. Others favored the tablet because they were accustomed to using tablets in everyday life, making the interface feel intuitive. Only one participant reported prior exposure to VR technology—having seen a grandchild use a gaming headset—which generated curiosity rather than apprehension. However, this limited familiarity did not necessarily translate into prior understanding of how visual field testing would function within a headset.

In summary, ease of use was closely tied to intuitive design, minimal setup requirements, and portability. Simple interaction, ease of instruction, device design and size/portability, and familiarity were the main subthemes. The VR and tablet devices were generally perceived as simpler and more adaptable, whereas the table-mounted structured setup was seen as both stabilizing for some and procedurally burdensome for others.

#### Psychological and motivational factors

Beyond physical and practical considerations, participants described psychological and motivational influences on their preferences. All participants articulated an overall evaluation of the devices. Many expressed a strong preference for VR, describing it as “definitely the best one,” while others favored the table-mounted due to familiarity or the tablet for its simplicity. Participants often weighed perceived comfort and clarity against their sense of trust in the device’s accuracy when forming their overall preference.

Novelty and enjoyment also contributed to motivation. Several participants described the VR as “fun” or compared it to a video game experience. This sense of novelty and engagement appeared to enhance willingness to complete the test and reduced perceived stress. For some, interacting with newer technology fostered a more positive emotional experience.

A smaller number of participants raised considerations related to cost and practicality. They observed that large, table-mounted perimeters require dedicated space and may be more expensive, whereas portable devices appeared more practical for smaller clinics or community settings. Although these comments were less frequent, they reflected an awareness of logistical and structural factors influencing device implementation.

In summary, psychological factors, including engagement, novelty, and perceived modernity, enhanced acceptance of portable devices. Subjective preference/overall evaluation, novelty and fun/emotional engagement, and cost and practicality were the main subthemes. Participants expressed greater enthusiasm toward technologies that felt less intimidating and more aligned with contemporary digital experiences.

## Discussion

This mixed methods study evaluated patient preferences for three visual field (VF) devices—the table-mounted Humphrey Field Analyzer (HFA), a tablet-based Melbourne Rapid Fields (MRF) system, and the virtual reality (VR)-based VisuALL headset—in medically underserved rural telemedicine settings. Our study showed a clear overall preference for portable devices, particularly the VR-based perimeter.

This study extends prior work on patient experience with visual field testing by demonstrating that the same usability and comfort advantages reported in tertiary settings and experimental cohorts also apply in medically underserved, community-based environments. Our findings suggest that when diagnostic performance is comparable, patient-centered attributes such as ergonomics, perceived simplicity, and engagement become decisive factors shaping technology acceptance and sustained use in real-world glaucoma care. These findings provide insight into how portable perimetry may enhance accessibility and patient-centered glaucoma monitoring in resource-limited environments.

More than half of participants preferred the VR-based platform for future testing, followed by tablet-based device, with the table-mounted being the least preferred. This mirrors clinic-based studies in which participants rated VR visual field paradigms as more attractive and enjoyable than screen-based and table-mounted perimeters, despite similar performance.[35, 36] In a VR oculomotor screening study, patients and controls judged the VR device as the most appealing modality compared to screen-based eye tracker and clinical standard table-mounted, underscoring the salience of user experience when multiple technically adequate options are available.[36] Similarly, a study using VR headsets with real-time eye tracking for remote visual field testing found high comfort and acceptance ratings, although many participants expressed a desire for some level of professional oversight.[37, 38] Our qualitative data echo these themes: participants emphasized comfort, reduced physical constraint, and intuitive interaction as reasons for preferring portable devices, while a few continue to value the perceived reliability and familiarity of table-mounted devices.

Ergonomic considerations emerged as a major determinant of preference and were particularly relevant in rural and aging populations. Participants described the chinrest, rigid posture, and dark-room requirement of the table-mounted perimeter as physically taxing, especially for those with neck or back problems, whereas both VR and tablet platforms permitted natural sitting positions, minor head movement, and use in standard exam rooms.[8, 21, 23, 39] Similar advantages of VR perimetry, such as smaller space take-up, elimination of eye patching, the possibility of testing with both eyes opened with their habitual glasses, and greater physical comfort, have been highlighted in clinical case reports and expert commentary. One case report described a successful VR-based field assessment in a previously non-assessable eye with severe glaucoma by leveraging binocular testing and fellow-eye fixation, illustrating how VR systems can extend functional testing to patients who struggle with clinical standard perimeters.[40] Importantly, comfort was not universally defined as freedom alone. Some participants expressed appreciation for the structured stability of the table-mounted, suggesting that ergonomic support and familiarity can mitigate discomfort. Nonetheless, the broader pattern indicates that reduced physical constraint and procedural simplicity enhance user acceptability. In our cohort, ergonomic freedom and absence of rigid support were framed as enabling, not merely convenient, particularly for older adults and those with mobility limitations—groups that often face compounded access barriers in rural settings.[8]

Participants highlighted the visual environment as a key influence on preference. The immersive visual environment provided by the VR headset also shaped patient experience. Participants in our study reported that blocking ambient light and peripheral visual distractions improved concentration and made the test feel more constrained and manageable. This controlled visual containment contrasted with the open peripheral awareness of the tablet and table-mounted perimeters.

This aligns with broader VR perimetry studies, which note that the head-mounted displays offer precise control over luminance and visual context and may enhance focus and reduce distraction.[8, 21, 22, 41] Avoiding monocular patching was frequently perceived as more natural and less disorienting, and binocular viewing was described as reassuring, a feature also emphasized by clinicians using VR devices in pediatric and glaucoma populations.[41] For many participants, this contributed to a sense of ease and confidence during testing. While some individuals seemed to value the familiar optics of the table-mounted device, the immersive qualities of VR headset were generally regarded as beneficial for maintaining attention and reducing distraction. Together, these observations suggest that immersive, distraction-limited environments may improve not only subjective comfort but potentially test reliability, although this requires further evaluation.

Perceived simplicity and cognitive load strongly influenced device adoption. Participants described both VR and tablet devices as straightforward, often requiring only understanding of the response button and minimal alignment, whereas the table-mounted device was associated with complex setup, such as trial lens placement, chin and forehead positioning, and greater uncertainty about “doing the test correctly.”[14] Reduced setup complexity and guided prompts lowered cognitive burden compared to the table-mounted alignment and patching procedures.

These findings align with the Technology Acceptance Model (TAM), which posits that perceived ease of use and perceived usefulness are central drivers of technology adoption.[42, 43] Participants’ emphasis on intuitive design, comfort, and procedural efficiency reflects how usability directly shapes acceptance of portable perimetry within clinical workflows. Prior studies have likewise reported that VR-based tests are seen as more intuitive and less cognitively demanding than the clinical standard table-mounted perimeter, particularly when they incorporate clear on-screen instructions and simple interaction measures.[21, 38] Familiarity also shaped perceptions of ease. Some participants preferred the table-mounted because of prior exposure, while others gravitated toward the tablet because it mirrored everyday device use, reinforcing the importance of prior technology experience in shaping ease-of-use perceptions.

Beyond usability, psychological engagement further differentiated device experiences. Several participants described the VR test as “game-like” or enjoyable, which reduced anxiety and increased motivation to complete the testing and potentially return for follow-up care. This is consistent with findings that VR-based visual tasks are rated more positively across user-experience dimensions than the clinical standard table-mounted perimeter, and with Self-Determination Theory (SDT), which posits that intrinsic enjoyment and perceived competence support sustained engagement.[35, 44, 45] In our data, the combination of intuitive interfaces, comfortable posture, and immersive environments appeared to bolster feelings of competence and autonomy—core SDT needs—potentially translating into better attention and fewer lapses during testing. At the same time, novelty and enjoyment were not sufficient on their own, some participants emphasized trust in accuracy and the reassurance of established technology, which resonates with a qualitative study in the UK on visual field testing on glaucoma monitoring, underscoring that innovation must be accompanied by clear evidence of clinical validity and effective communication about comparability to the clinical standard.[14, 38]

This balance between innovation and credibility highlights the need for portable technologies to demonstrate not only usability but also clinical reliability. Collectively, these findings resonate with patient-centered care principles, which emphasize how structural features (device design, ergonomics, portability) and process factors (testing experience, clarity of instructions, comfort) influence patient satisfaction and engagement. Within this framework, portable perimetry improves both the structural accessibility and experiential quality of glaucoma monitoring. The alignment with TAM and SDT further suggests that enhancing perceived ease of use and practical utility may accelerate the adoption of portable technologies in ophthalmic telemedicine programs.

The findings suggest that portable perimetry may address structural and experiential barriers to glaucoma monitoring in underserved communities. These preference patterns have important implications for telemedicine and home-based glaucoma monitoring. Studies of remote VR visual field testing have shown that patients generally accept and tolerate headset-based assessments and appreciate the potential to test outside the hospital, but many still express a desire for some degree of supervision or support, whether in person or remotely.[37] Our findings are concordant: while participants welcomed the convenience and comfort of portable devices in FQHCs, they also valued guidance from staff and the reassurance of being “looked after.” This suggests that ophthalmic telemedicine models using portable perimetry in underserved settings should pair decentralization of the device with maintained relational and informational support, such as remote coaching, periodic in-person review, rather than assuming that portability alone guarantees sustained engagement. Importantly, recent validation and systematic review work indicate that multiple VR and tablet-based platforms can achieve diagnostic performance comparable to table-mounted for visual field parameters, supporting the feasibility of substituting or complementing the clinical standard perimeter where appropriate.[20–23, 39, 41, 46] Emphasizing this equivalence to patients is important to maintaining perceived usefulness when transitioning from traditional to portable modalities.[14]

Finally, our findings reinforce that patient preferences are not merely a secondary consideration but a structural determinant of long-term adherence in disease management. Participants emphasized that shorter, more comfortable tests would increase their willingness to return for follow-up visits. In underserved rural areas, where travel, time off work, and clinic capacity already limit access, platforms that are more comfortable, quicker, and deployable in primary care or community settings may substantially reduce drop-out and missed visual field monitoring. The literature suggests that patients are willing to tolerate discomfort when they believe a test is essential, but they consistently favor modalities that minimize physical and cognitive burden when reliability is maintained.[14, 35, 37, 38] Our study extends this observation to FQHC-based telemedicine, indicating that integrating validated portable perimetry into these programs can simultaneously address infrastructural constraints and align with patient preferences. Future work should move beyond cross-sectional preference assessment to evaluate whether the use of portable devices in such settings improves longitudinal adherence, test reliability over time, and ultimately clinical outcomes, while also incorporating clinician perspectives and cost-effectiveness analyses to guide scalable implementation.

In conclusion, this mixed methods study demonstrates that patients in medically underserved rural areas prefer portable visual field devices, particularly VR-based, over conventional table-mounted perimetry. Comfort, ergonomic flexibility, immersive visual experience, simplicity of interaction, and motivational engagement emerged as key determinants of preference. Portable perimetry offers a promising approach to expanding glaucoma screening and monitoring within ophthalmic telemedicine frameworks and community-based care models.

## Data Availability

All relevant data are within the manuscript and its Supporting Information files.

## Acknowledgments

The authors thank the research coordinators, participants, and clinical staff of the federally qualified health centers involved in the AL-SIGHT program for their support and collaboration. EKAA gratefully acknowledges support from the American Association of University Women (AAUW) through the International Doctoral Fellowship.

## Notes

### Competing Interest Statement

I have read the journal's policy and the authors of this manuscript have the following disclosures: E.K. Antwi-Adjei, None S. Datta, None C.A. Girkin, Heidelberg Engineering (F), Topcon (F) C. Owsley, Johnson & Johnson (C), Sanofi (C) L.A. Rhodes, IdentifEye Health (C) M. Fifolt, None L. Racette, Olleyes, Inc. (C). These relationships do not alter our adherence to PLOS ONE policies on sharing data and materials.

### Author Declarations

The protocol was approved by the Institutional Review Board of the University of Alabama at Birmingham, and all procedures adhered to the tenets of the Declaration of Helsinki.

